# Th2 infiltration is a better predictor of survival than tumor-infiltrating lymphocytes (TILs) in triple-negative breast cancer (TNBC)

**DOI:** 10.1101/2023.06.02.23289891

**Authors:** Susie Brousse, Florence Godey, Patrick Tas, Boris Campillo-Gimenez, Elodie Lafont, Amanda Poissonnier, Jean Levêque, Vincent Lavoué, Matthieu Le Gallo

**Affiliations:** Centre de lutte contre le cancer Eugène Marquis, Rennes, France; Inserm UMR_S 1242, Chemistry Oncogenesis Stress Signaling, University of Rennes; Department of Pathology, Rennes University Hospital, Rennes, France; Inserm, LTSI - UMR 1099, F-35000 Rennes, France; Department of Gynecology and Obstetrics, Rennes University Hospital, Rennes, France

**Keywords:** Triple-negative breast cancers, tumor-infiltrating lymphocytes, immune response, Th2 signature, prognostic and predictive factors

## Abstract

**Purpose:** Triple-negative breast cancers (TNBC) account for 15% of all breast cancers but carry the worst prognosis. Because of their heterogenicity, these tumors are not all prone to targeted therapies. However, due to their high immune infiltration, targeting their immune microenvironment is of tremendous interest and is becoming the standard of care for high-risk early-stage TNBC. Nevertheless, the characterization of this immune infiltrate is often limited to general tumor-infiltrating lymphocytes (TILs) counting, without characterization of lymphocytes subtypes. Thus, we aimed at precisely characterizing these sub-populations and evaluating their prognostic significance.

**Methods:** We selected 91 TNBC tumors for which we had both the TILs count on hematoxylin and eosin (H&E) slides determined by an expert pathologist and the immune microenvironment cell subtypes characterization using flow cytometry (FC). We then compared the prognostic value of immune microenvironment subpopulations vs total TILs count.

**Results:** TNBCs contained a mean of 22.8±25.9% TILs in the tumor surface area, including mainly CD4+ helper T lymphocytes (14.1%), mostly Th2 (11.7%), CD8+ cytotoxic T lymphocytes (11.1%), and myeloid cells (8.4%) including antigen presenting cells (APC). The TILs count was correlated with the abundance of these cellular subpopulations (p≤0.004). TILs percentage was predictive of overall survival (OS) in univariate analysis (p=0.044), high APC infiltration was predictive of relapse-free survival (RFS) in univariate analysis (p≤0.030), and Th2 infiltration was predictive of both RFS and OS in univariate (p=0.009, 0.008 respectively) and multivariate analyses (p=0.002, 0.010 respectively).

**Conclusion:** The characterization of TILs composition is essential to better understand the potential antitumoral functions of these cells, and to strongly improve the associated prognostic and predictive values. We here demonstrate that Th2 subpopulation is associated with a better overall survival in TNBC and could be of use to predict response to the newly used immunotherapies.

## Introduction

Representing 25% of newly diagnosed cancers among women, breast cancer is a worldwide health concern with 685,000 associated deaths in 2020 according to the World Health Organization, making this pathology the leading cause of female death by cancer in most countries (1,2). Triple negative breast cancers (TNBC) are defined as negative for the expression of hormonal receptors (HR) to estrogen (ER), progesterone (PR), and Human Epidermal Growth Factor 2 (Her2) receptors, being mostly constituted of basal-like molecular subtype (3,4). Whilst they represent 10 to 20% of all breast tumors (2,5), they are a major health concern due to their epidemiology, being frequently diagnosed among younger non-menopausal women, and to their quickly unfavorable evolution (6–8) associated with a high proliferation index, frequent relapse after chemotherapy (9,10), and early metastatic disease (pulmonary, hepatic, neurologic) (8,11). Besides, we have too few clinically available targeted therapy (5,9) since they represent a very heterogenous disease made of up to 7 subtypes (12–14).

Even though there are some promising therapeutic options (12,13,15–18), TNBC do not share a common antigenic target, making it difficult to find broadly efficient therapies. Nevertheless, their immunogenicity could offer some therapeutic options, since an immune infiltrate is detected in up to 75% of TNBC, much more than in luminal subtypes (19), and its composition is very distinct from the other molecular subtypes (19,20). Hence, immunotherapy recently became available and recommended for immunosuppressive TNBC, thanks to the the newly FDA-approved Atezolizumab (21) and Pembrolizumab (22), sometimes depending on PD-L1 expression.

However, PD1/PD-L1 expression does not select responding patients accurately enough (22,23). So, in order to better identify which patients could benefit from immunotherapies and to identify new potential targets, it is essential to better understand the role and composition of the immune infiltrate in TNBCs.

The tumor-infiltrating lymphocytes (TILs) density has been shown to be predictive of response to neoadjuvant chemotherapy (NAC) and of patient outcome (24–26), especially in TNBC (24,25,27,28). CD8+ cytotoxic T cells (TC) were first proved to be markers of good outcome, especially in TNBCs (26,29,30), as well as CD4+ helper and CD4+ follicular helper (fh) TC (31,32). But TNBC can escape immune cell-mediated cytotoxicity by developing an immunotolerance thanks to immune checkpoints and regulatory TC (Treg) activation (33). Moreover, the modulation of the expression of some immunological-related genes (34), and different immune signatures such as T helper cell type 1 (35), low M2-like macrophages (36) and gene cluster Immunity 2 (37) also have prognostic value. That’s why further classification in immunocompetent or tolerant TC is necessary to evaluate their prognostic significance.

Nevertheless, to assess immune infiltrate in daily practice, pathologists currently only measure TILs density (38,39), since it is predictive of pathological complete response (pCR) to NAC, and of significantly better recurrence-free (RFS) and overall survival (OS) (40). However, no therapeutic option derives from this measurement since the TILs subtypes are not searched for. Hence, we here aim to characterize the immune microenvironment of TNBC tumors by flow cytometry and thus to better specify the prognostic value of TILs subtypes.

## Methods

### Patients

123 TNBC patients were recruited at the time of surgery, before any chemotherapy and radiotherapy was administered, from January 2013 to December 2018 in Centre Eugène Marquis in Rennes, France. All participants provided informed consent and the study was approved by the local Ethics Committee Review Board and complied with local ethics guidelines. 91 patients were finally selected (Supplementary Figure 1).

### Sample collection

After surgical resection, TNBC tumor fresh tissue was taken by the pathologist in the operative room and processed into two fragments. The first one, fixed in 10% neutral buffered formalin for standard histochemical analysis with Hematoxylin and Eosin (H&E), was analyzed for TILs quantification and requalified for research purposes. The second one was cut into small pieces (<2 mm), stored in cold RPMI medium (Gibco), and processed within two hours with Human tumor dissociation kit (Miltenyi Biotec GmbH) in a gentle MACS Dissociator, according to the manufacturer’s recommendations. Macroscopic pieces were eliminated using a Corning cell strainer (70 μm), dissociated cells were then washed twice in RPMI (20 mL) and counted using a hemocytometer. Living cells were immediately analyzed using multiparameter flow cytometry (FC).

### Analyses of TILs by FC

Tumor cells (50,000 cells) were suspended in PBS supplemented with 2% BSA, 2% FCS, and FcR block (Miltenyi Biotec GmbH®) at 4°C for 20 minutes. Cells were then stained for 30 minutes at 4°C with fluorochrome-coupled antibodies targeting CD45-PeVio770 (clone 5B1), CD3-FITC (clone REA613), CD4-APCVio770 (clone VIT4), CD8-FITC (clone BW135/80), CD11c-APC-Vio770 (clone REA618), CD83-APC (clone HB15), CD86-FITC (cloneFM95), CXCR3-PE (clone REA232), CCR6-APC (clone REA190) (Miltenyi Biotec GmbH), CD14-APC (clone MφP9), CD24-PE (clone ML5), CD44-APC (clone M44-26) (BD Biosciences, France). Matching isotypic antibodies purchased from the same manufacturers were used as control. Cells were then washed twice in PBS supplemented with 2% BSA and 2% FCS and resuspended in PBS. To assess cell viability, cells were incubated with 7-AAD (BD Biosciences) for 10 min prior to cytometry analysis. Data were acquired using a FACS Canto II (BD Biosciences) or a Novocyte cytometer (ACEA Biosciences) and analyzed using Novoexpress software. Tumor and immune cells were defined as described in Table 1 and Supplementary Figure 2. The different TILs populations were thus measured, and their abundance, overall and relative over total CD45+ immune cells, was quantified for each tumor (Figure 1).

**Figure 1-.**
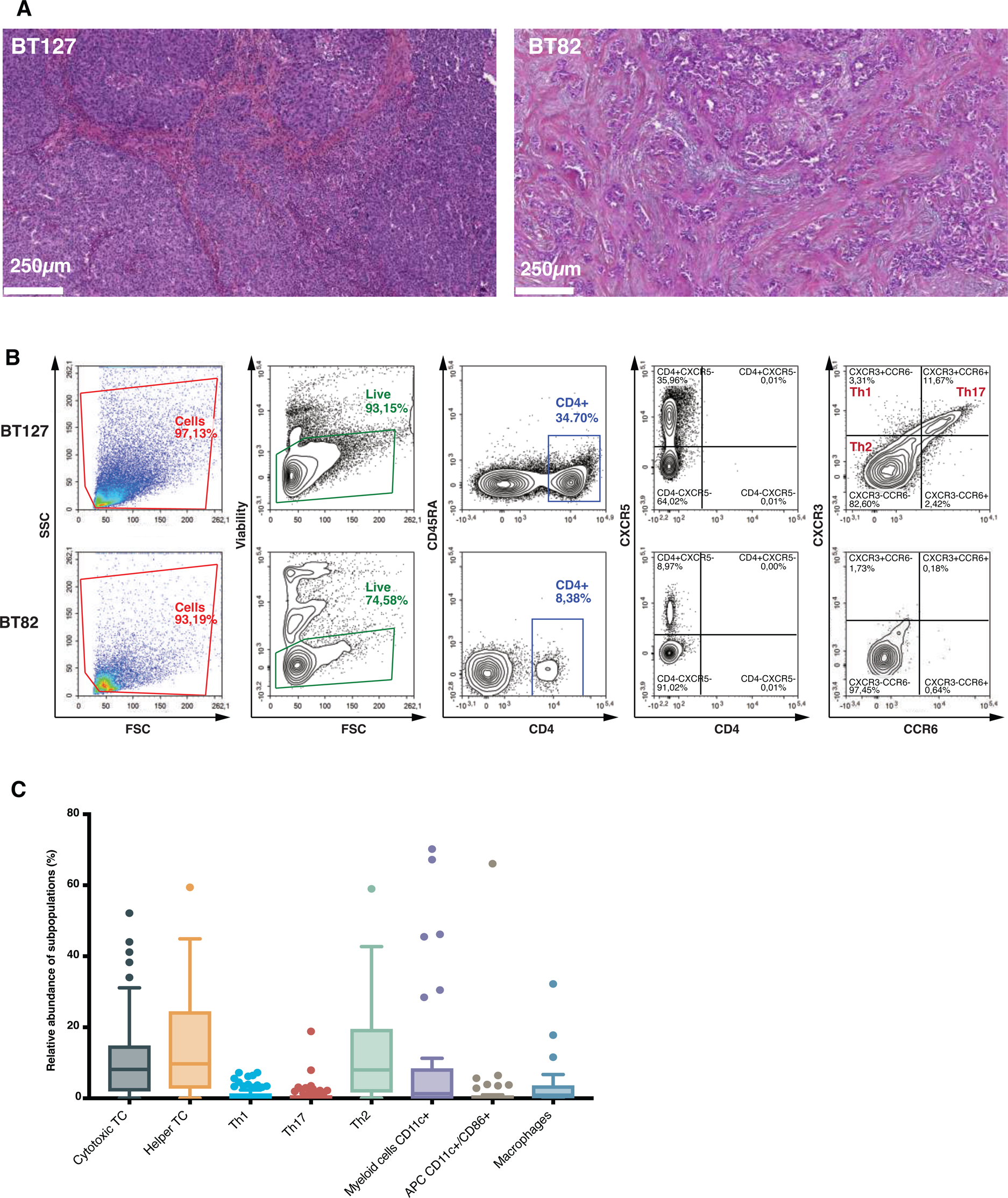
Tumor-Infiltrating Lymphocytes (TILs) and their characterization by Flow Cytometry (FC). Corresponding H&E pathological slides (A) and FC analysis (B) of a highly infiltrated group C tumor, BT127, showing 70% of TILs infiltration (A) and 20% of Th2/CD45 +cells (B), and a poorly infiltrated group A one, BT82, showing 5% of TILs stromal infiltration (A), and 6% of Th2/CD45+ cells (B). TILs count consists of mononuclear cells count in stromal fibrous spans (pink). Th2 cells are defined as living T helper lymphocytes (CD4+) expressing neither CXCR3 (contrary to Th1 and Th17 lymphocytes) nor CCR6 (contrary to Th17 lymphocytes). C: Complete characterization of the immune microenvironment of all tumors by FC showing the relative abundance of the different subpopulations (Percentage values in Supplementary Table 3). TC: T lymphocyte cells; APC: Antigen-presenting cells.

**Table 1-.**
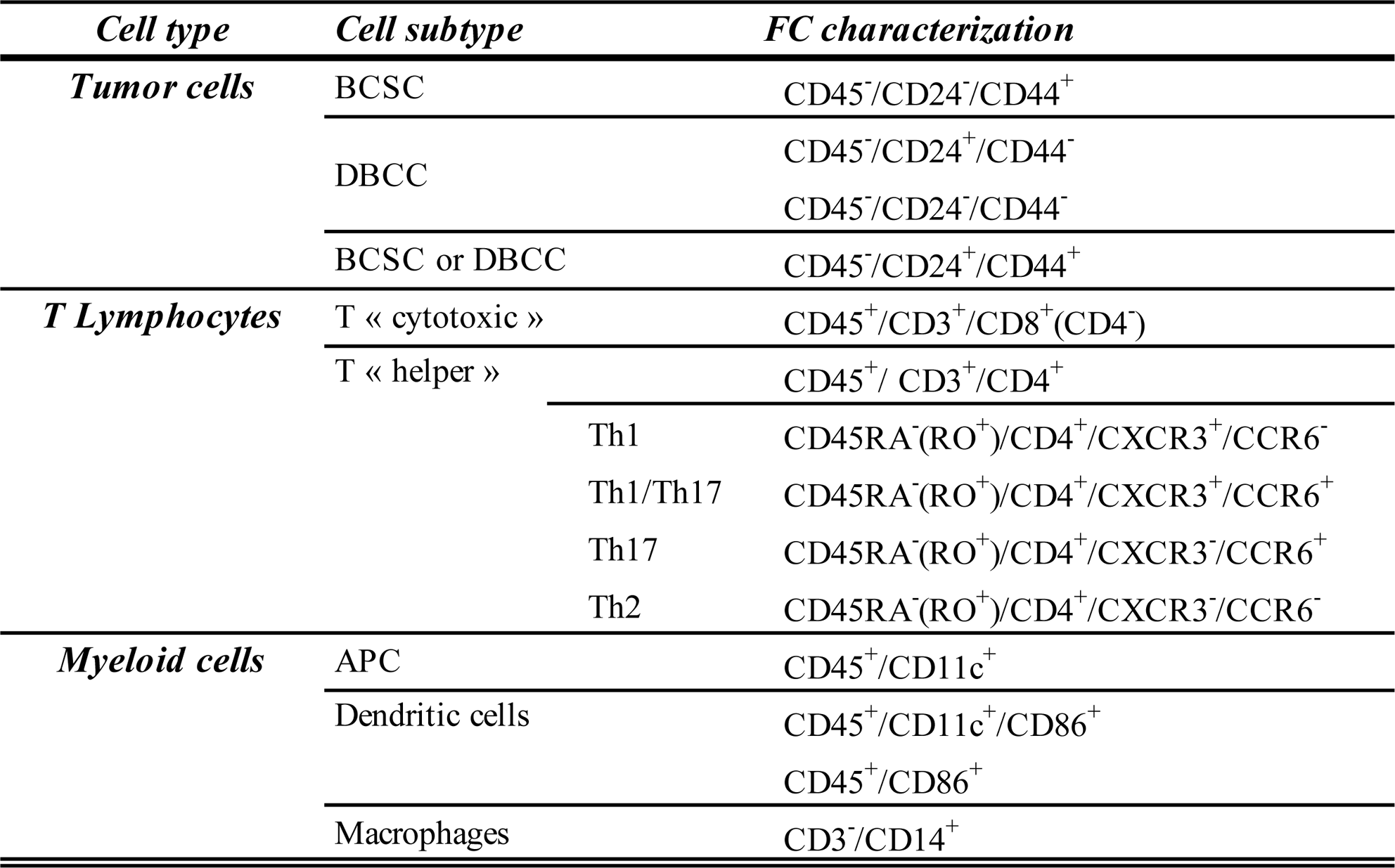
TILs characterization by Flow Cytometry (FC) according to the expression of their surface markers. BCSC: Breast Cancer Stem Cells; DBCC: Differentiated Breast Cancer Cells; APC: Antigen-Presenting Cells.

### TILs quantification

H&E FFPE slides were obtained from Ouest pathology after pathological report was done. Retrospective TILs count, defined as the percentage of tumor stroma surface occupied by TILs, was done based on the 2014 TILs working group guidelines updated in 2018 (38,39). Group A, B, and C were defined as TILs infiltration <20%, ≥20% and <50%, and ≥ 50% respectively (Figure 1).

### Statistical analyses

Patients and tumor characteristics were compared between TILs or Th2 groups using Chi-squared tests or Fisher’s exact tests for qualitative variables, and either Student’s t tests, Mann Whitney U tests, or ANOVA and Kruskall-Wallis tests (when more than 2 groups were compared) for quantitative ones. Pearson or Spearman correlations were used to study the link between TILs percentage and other immune subpopulations abundance. Relapse-free survival (RFS) was defined as the time from surgery to relapse or death and overall survival (OS) as the time from diagnosis to death. Patients lost to follow-up were censored at the date of last news. Kaplan-Meier curves were estimated for RFS and OS, and log-rank tests were performed to compare survival distributions. Univariate and multivariate Cox regression analyses were performed to calculate unadjusted and adjusted Hazard Ratios (HR) and were presented with 95% Confidence Intervals (CI95%). Bayesian Information Criterion (BIC) was used to select independent parameters according to a forward step-by-step selection procedure. Whole statistical analysis was performed with R and RStudio® softwares. All reported p values were two-sided and considered statistically significant when <0.05.

## Results

### Population description

91 patients got a complete immune characterization (TILs count by pathologists and subpopulations characterization by FC) (Supplementary Figure 1). Their characteristics are precisely described in Supplementary Table 1 and 2. Briefly, our population consisted of females with a mean age of 58 (±14.5) years. Of these women, 15.7% and 11.8% carried a BRCA1 or BRCA2 mutation respectively. Their tumors were mostly high grade (93.4%) and highly proliferative (mean Ki67: 64.8±23.8%) invasive ductal carcinomas (85.7%) of 22.9±11.6mm with 1.5±3.6 metastatic lymph nodes. Therapy mostly consisted of lumpectomy or breast conserving surgery (75.8%), with axillary treatment (94.4%), followed by chemotherapy (85.7%), and radiotherapy (86.5%). The median follow-up was 28.0 months (Supplementary Table 2).

### TILs count, characterization using FC, and their correlation

Most TNBC (58.2%) were poorly infiltrated Group A tumors, versus 23.1% Group B and 18.7% Group C, for a TILs count of 22.8±25.9% (median 10.0% [0.0-90.0]). TILs consisted of 3.1% macrophages, 8.4% myeloid cells, 11.1% CD8+ cytotoxic TC, and 14.1% CD4+ helper TC (1.0% Th1, 0.8% Th17, and 11.7% Th2 subsets) (Figure 1 and Supplementary Table 3).

The correlation between TILs groups and immune cells measured by FC was linear and significant for total leukocytes (p≤0.001), helper TC (p≤0.001), cytotoxic TC (p≤0.001), Th17 and Th2 responses (p≤0.01), and antigen presenting cells including dendritic ones (p≤0.001 & p≤0.01 respectively) (Figure 2). Pearson and Spearman correlations showed the same results when considering TILs percentages and total leukocytes (p=0.002), myeloid cells (p<0.001), cytotoxic TC (p=0.002), helper TC (p<0.001), and Th17 (p=0.022) or Th2 expressing TCs (p=0.002). We noted that the presence of Th1 TCs was not significantly correlated with TILs group (p=0.14) or percentage (p=0.218).

**Figure 2-.**
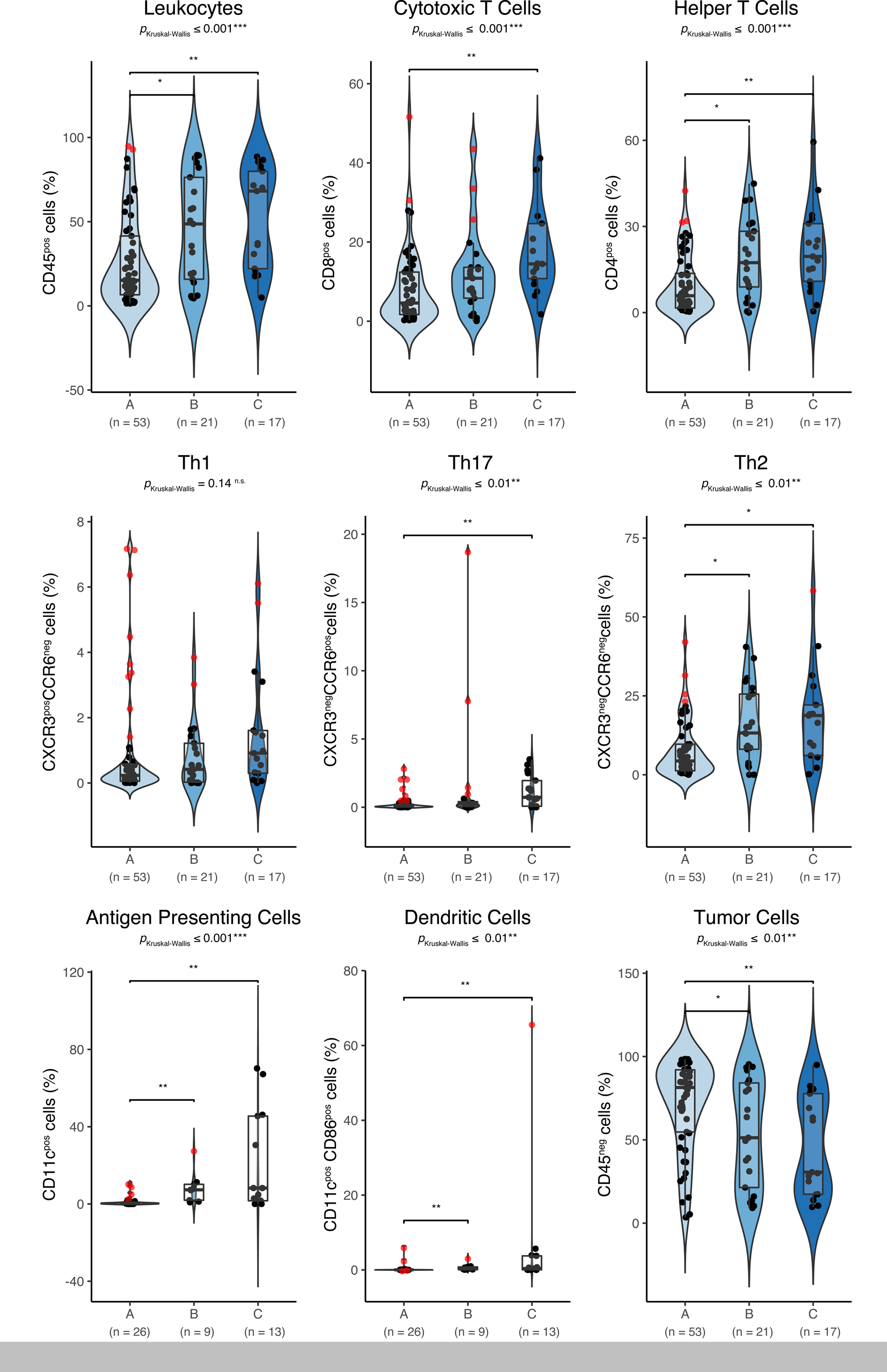
Correlation of TILs groups with immune subpopulations relative abundance. Correlation of TILs group as defined by the pathologist with total leukocytes, including helper CD4+ and cytotoxic CD8+ T cells, antigen-presenting cells (APC) or dendritic cells, and tumor cells characterized by flow cytometry (FC). Helper T cells could either trigger a Th1, Th17, or Th2 response. Only populations with significant correlation are shown on the figure. p values were derived from a Holm and a Kruskal-Wallis tests (for the comparison of respectively 2 and 3 groups). ns: non-significant; * p≤ 0.05; ** p≤ 0.005; *** p≤ 0.001.

### Correlation of demographical and tumoral characteristics with immune cells

TILs groups and percentages were not significantly correlated to patients’ clinical characteristics, except for BMI (p=0.038 and 0.007 respectively) and the presence of BRCA mutations (p=0.009 and 0.016 respectively). Regarding tumor characteristics, only DCIS (p=0.037) and grade (p=1.8.10^−9^) were correlated with TILs percentages. Th2 response was only significantly related to intravascular tumor emboli (p=0.047) and there was a trend of an association with both lymph node involvement and grade (p=0.123 and 0.180 respectively) (Supplementary Table 4).

### RFS and OS analyses

16 patients (17.6%) relapsed, mostly by distant metastasis (15.4%), and 10 (11%) died of cancer-related cause resulting in a 5-year RFS and OS of 77% and 83% respectively (Supplementary Table 2 and Supplementary Figure 3). Only BMI was significantly predictive of relapse and OS (p=0.033 and 0.034 respectively) (Table 2). Number of foci and pathological size were significantly predictive of OS (p=0.009 and 0.010 respectively). Pathological size and nodal involvement were also strongly predictive of both RFS and OS in multivariate analyses (p<0.05)(Table 2 and Figure 3). All the non-significative variables are shown in Supplementary table 5.

**Figure 3-.**
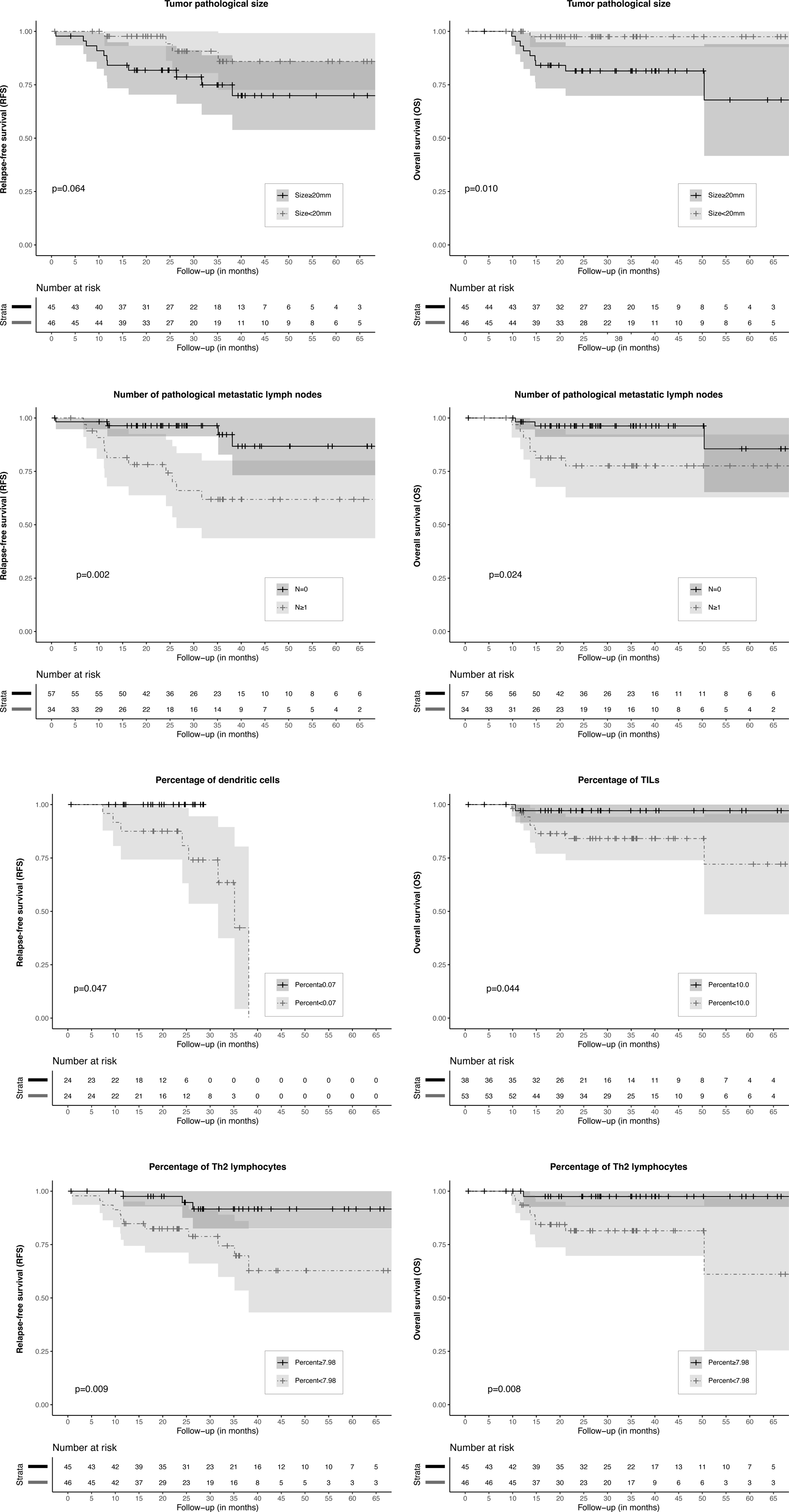
Relapse-free (RFS) and overall survival (OS) analyses. RFS and OS according to tumor pathological size (median=20mm), pathologically confirmed lymph node metastasis, dendritic cells percentage by FC (median=0.07%), tumor-infiltrating lymphocytes (TILs) percentage (median =10.0%), and Th2 lymphocytes percentage by FC (median=7.98%). Kaplan Meier curves were estimated, log rank tests (for p values) were performed to compare survival distributions. p values in bold font are significant ones.

**Table 2-.**
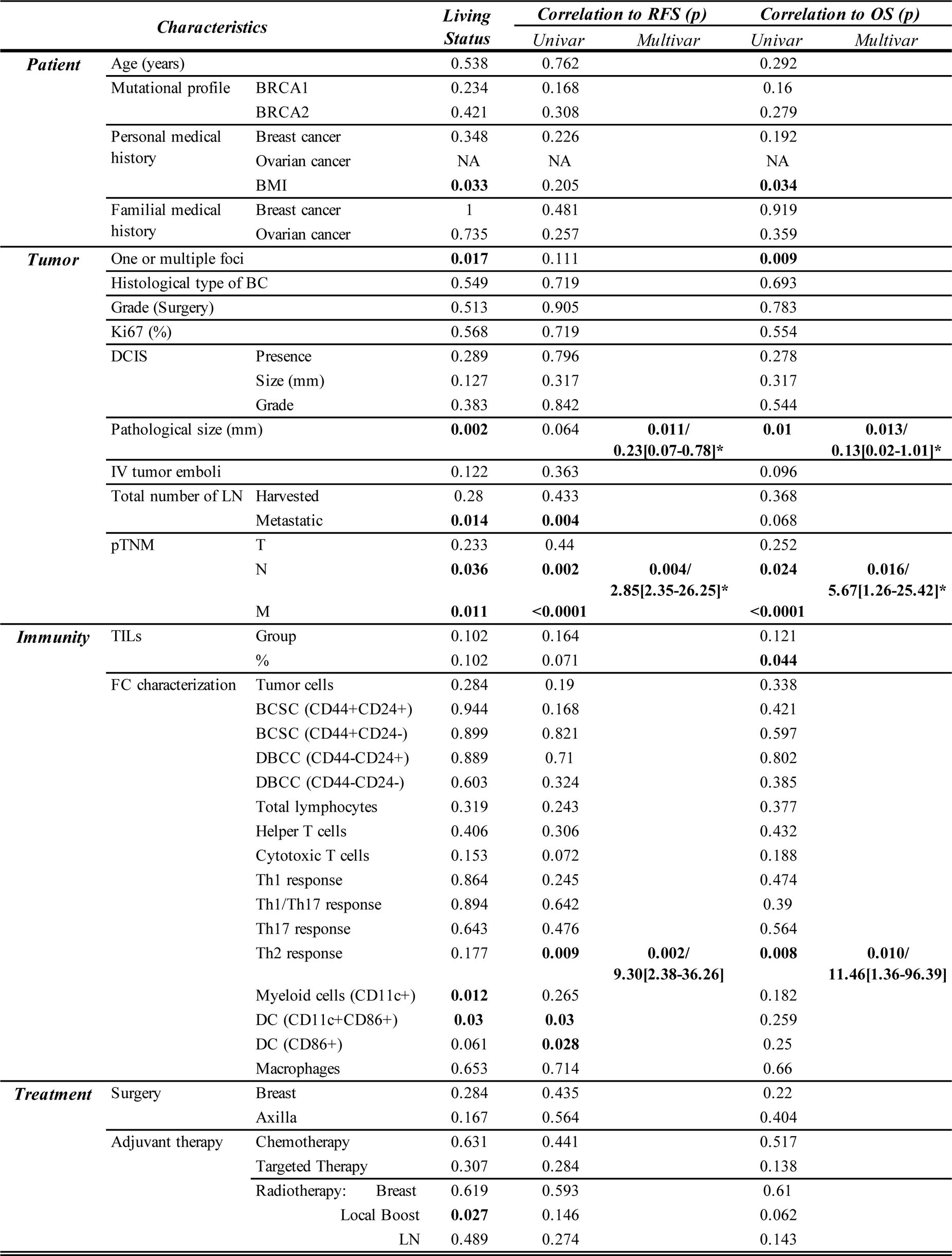
Correlation of patient, tumor, immunity, and treatment characteristics to relapse-free survival (RFS) and overall survival (OS). p values for survival status are obtained using Wilcoxon test for quantitative variables and Chi2 or Fisher exact test for qualitative ones, depending if the group do or do not contain at least 5 values respectively. p values for relapse free and survival regression are obtained using log rank test for univariate (univar) analyses and Cox model for multivariate ones (multivar), using BIC condition for prognostic usefulness. p values <0.05 appear in bold font in the text. *p values/ HR [CI95%]. BMI : Body mass index; DCIS : Ductal carcinoma in-situ; IV : Intra-vascular; TILs : Tumor-infiltrating lymphocytes; FC : Flow cytometry; BCSC : Breast Cancer Stem Cells; DBCC : Differentiated Breast Cancer Cells;DC : Dendritic Cells; pTNM : pathological staging of tumor size(T), lymph node involvement(N), and metastatic status(M) of breast cancer according to international guidelines.

The TILs percentage was predictive of OS (p=0.044) (Figure 3 and Table 2) but only tended to be of RFS (p=0.071). The percentages of CD45+ leukocytes, CD4+ helper TC, and macrophages were not significantly predictive of RFS or OS, and we observed a non-significant trend with CD8+ cytotoxic TCs (p=0.072 and 0.188 respectively). However, we discovered that a high Th2 infiltration was strongly predictive of both RFS and OS (Figure 3) in univariate (p=0.009 and 0.008 respectively) and multivariate (p=0.002 and 0.010 respectively) analyses (Table 2), while Th1 and Th17 were not. Myeloid cells including dendritic ones were also significantly predictive of relapse (p=0.012 and 0.030 respectively) (Table 2).

When considering the different Th populations, only Th2 was a better predictive factor of survival, especially OS, than total leukocytes (Figure 4). In accord, almost all patients having relapsed or died were Th2 low (below the median), even though some of them where highly infiltrated by lymphocytes. Th1 and Th17 did not add the same prognostic value, and Th17 was particularly poorly correlated to RFS and OS (Figure 4).

**Figure 4-.**
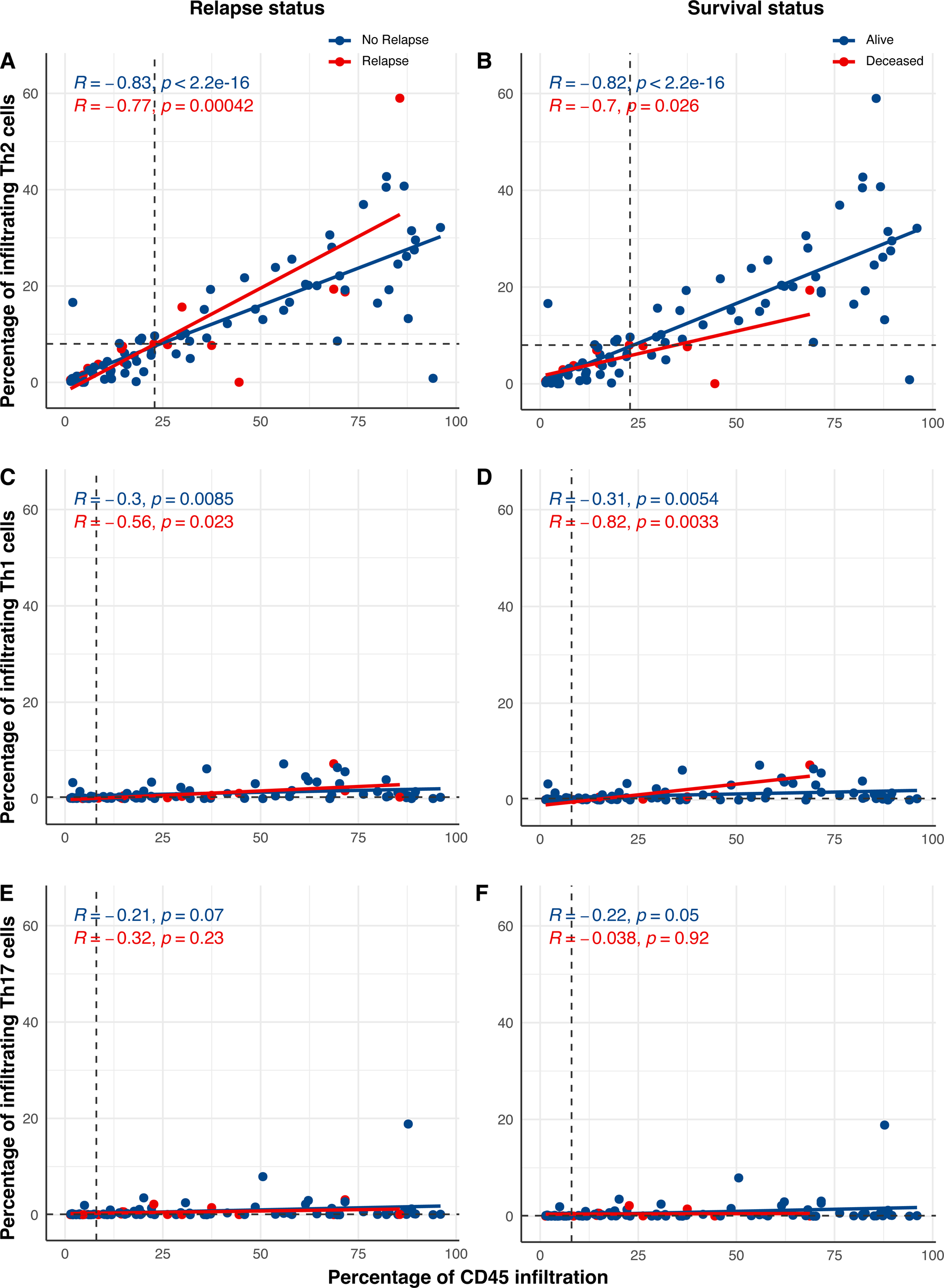
Relapse and overall survival status of patients depending on Th subpopulation versus total CD45+ leukocytes infiltration measured by flow cytometry (FC). Repartition of free from relapse (no relapse in blue) patients versus having-relapsed ones (relapse in red) (A,C,E) and alive (red) versus deceased (blue) ones (B,D,F) ones according to Th2(A,B), Th1 (C,D), Th17 (E,F) infiltrations vs all leukocytes. Dashed lines represent medians of different subpopulations infiltrations.

## Discussion

The emergence of immunotherapy represents a new hope for TNBC patients since these tumors are frequently highly infiltrated by immune cells and may thus respond to immune checkpoint inhibitors. However, markers than can robustly predict responsiveness to immune checkpoint therapy are lacking in TNBC. Furthermore, the nature of this immune infiltrate remains largely undocumented. Hence, it is essential to better characterize it to identify patients that would benefit the most from these new therapies.

### TILs content, methods of investigation, limits, and interpretation

TILs are needed to recognize disseminated tumor cells or death-associated molecular patterns (DAMPs) to trigger further immune recruitment, infiltration, and activation of immunogenic cancer cell death (41). Regardless of their localization, the characterization of TILs subtypes is critical, since they can have either a pro-or an anti-tumorigenic function (42). Memory, helper, or CD4^+^ fh TC can modulate the immune response, while cytotoxic CD8^+^ TC play a main role in cancer killing. FOXP3+ Treg or the activation of immune checkpoints (PD1/PD-L1) and CTLA4 are crucial in immune tolerance (33). Immune escape (drop in CD8+ and CD4+ TC) and tolerance (by PD-L1, PD-L2, and HLA-1 overexpression (43) or Treg activation (44)) are associated with poor prognosis and with the development of metastatic disease. But results are still controversial (29,45–48), also depending on tumoral TILs localization (49). Zhang et al. even showed that Treg and PD-1/PD-L1 expression was correlated with better OS in TNBC That is why immunomodulatory subtypes such as Treg and Th2 CD4+ TC need to be considered and better understood in TNBC.

TILs percentage is currently the only assessed parameter in the clinic but has many limits. First, there is a need for standardized TILs measurements by pathologists (38), since variability can affect the correlation of TILs with pCR (51). Moreover, the TILs working group only recommends the measurement of stromal TILs that are easier to measure than intratumoral ones (39,52). Nevertheless, results are still controversial (31), since Ruan et al. showed that both intratumoral and stromal TILs were independent predictors for pCR (28).

In our study, and contrary to the data previously mentioned (24–26), stromal TILs were not significantly associated with RFS and OS, perhaps because of the limited amount of relapses and death events in the studied cohort. However, other studies also failed to show a prognostic utility of TILs count alone (53). CD8+ and CD4+ TC were not significantly predictive of RFS or OS in our cohort either, while they frequently are in TNBC (24,26,29–32,48,54), maybe for the same reasons.

### Need for reproducible immune characterization

In our study, we counteracted these limitations by immunophenotyping the immune infiltrate on fresh tumor, without consideration of localization using a highly standardized and trustworthy FC method. Other studies use whole exome sequencing to assess T cell infiltration and correlate these data to prognosis and response to immunotherapy (55), or even gene-expression deconvolution to characterize T cell subtypes (53,56–58). However there is currently no consensual transcriptomic signature for Th2 cells.

### Th2 prognostic value

We observed that Th2 infiltration was the most frequent one, as previously reported (20). The prognostic value of Th2 in various cancers is often unconclusive or assumed to favor pro-tumorigenic immune-tolerance because of its link to aggressive features (59–65). On the contrary, high Th2 infiltration was a strong factor of good prognosis in our cohort, as already suspected in previous studies through thymic stromal lymphopoietin (TSLP) (66,67), or collaboration with eosinophils (68,69), sometimes being as powerfully antitumoral as Th1 (70,71). Th2 levels even favorably correlated to pCR in ER+ breast cancers (72). AllergoOncology group showed that immune escape can be reversed by IgE and Th2 response (69,73,74). Th2 anti-tumoral action could be boosted by anti-TGF-β (75) and Th2/Th1 response could be recapacitated by xanthohumol (76), saikosaponin (77), anti-IL4/IL4R (78), anti-IL13 (79), anti-CCR4 (80), among others (70,81). Other interesting perspectives consist of better understanding Th2 interaction with other immune subsets, like fibroblasts (82), macrophages (83,84), dendritic cells (79,85), or eosinophils and IgE (68,86), to better control Th2 response. Finally, immunotherapies modulating Th2 response could also be used around the time of surgery that has been proved to induce immunomodulation (87).

### DC prognostic value

We also showed that dendritic cells, potentially through their interaction with Th2 CD4+ TC, were correlated with better survival. On the contrary, DC have also been reported as immunosuppressive (79), especially through Th17 response (85). To face this complexity, it will be required to consider the DC subset involved (88). In addition, studying a wider signature including B-cells and myeloid-derived antigen-presenting cells (89), such as the ones used to predict response to treatment (90,91) will likely be informative. Of note, immune metagenes overexpression and innate and adoptive immune cooperation are also associated with low-risk of metastasis in TNBC (91,92).

### Clinical features and immune microenvironment

Only BMI correlated with TILs infiltration in our cohort, emphasizing the complex immune relation between obesity and cancer (93), especially through Th2 processes (94). Few of the tumoral characteristics were correlated to TILs infiltration, as previously reported (20). Nevertheless, Th2 percentages was significantly correlated with intravascular tumor emboli, a variable often linked to S.B.R. grade and lymph node involvement. Th2 levels were already proved to be higher in tumor microenvironment and lymph nodes in case of aggressive, therapy-resistant, metastatic, and large or locally advanced breast cancers (30,72,95–99), confirming their abundance without proving their pejorative prognostic value.

### Strength and weakness of our study and added value

Potential limitations of the applicability of our results could be the good RFS and OS of our patients compared to general TNBC populations (8), and the inclusion of patients who underwent surgery first whereas NAC is nowadays the standard of care for numerous TNBC. However, identifying a Th2 response in supposedly favorable-prognosis BC could be of tremendous interest to prevent unexpected but still early and deadly relapses. Moreover, even if TILs assessment post-NAC has some specificities (39,100), it is transposable to the ones performed pre-NAC (35,101), including for Th response in tumor (99) and in lymph nodes (95). TILs assessment post-NAC is also an independent predictor of survival (40,102), even better when used alongside Residual Cancer Burden score (RCB) (103). Besides, TILs count can also be done on pre-NAC biopsies and still carry the same prognostic value (104). Finally, modification of immune subpopulations percentages and ratios has also been proved to be of importance after NAC (47,105), and could thus be studied at both timepoints.

A better understanding of the TILs in TNBC, especially Th2 (75), could eventually enable to target them more specifically (106–108), or even to use them for Adoptive cell transfer (109), CAR-TC design, or even vaccines (21,110). However, we still need to identify which tumors are immunogenic enough to benefit from it (111,112). Following the CREATE-X trial, adjuvant chemotherapy (Capecitabine) is currently given to non-pCR TNBC patients regardless of the immune infiltrate (113), while only some of them will benefit from it. Hence, immune characterization could predict chemotherapy efficacy or immune exhaustion making relevant the addition of immunotherapy to Capecitabine for some selected non-pCR TNBCs, similarly to those benefiting from Atezolizumab in metastatic settings (21). Finally, necessary future therapeutic development include further understanding of the role of other immune cells such as macrophages (114), natural killer TC (NK cells) (115), and their interactions with TILs and Th2.

## Data Availability

All data produced in the present study are available upon reasonable request to the authors

## Abbreviations

BC: Breast Cancer
BCSC: Breast Cancer Stem Cell
CD: Cluster of Differentiation
DBCC: Differentiated Breast Cancer Cell
DC: Dendritic cells
DCIS: Ductal Carcinoma In Situ
FC: Flow Cytometry
FFPE: Formalin-Fixed Paraffin-Embedded
H&E: Hematoxylin and Eosin
LN: Lymph Node
NAC: Neoadjuvant chemotherapy
OS: Overall Survival
pCR: pathological Complete Response
pTNM: pathological classification of Tumor size (T), Node involvement (N), and Metastasis (M) according to international guidelines
RFS: Relapse-Free Survival
TC: T Cells (Lymphocytes)
Th response: T helper response
TILs: Tumor-Infiltrating Lymphocytes
TNBC: Triple-Negative Breast Cancer

## Acknowledgements

SB This work was funded by grants from Rennes University Hospital (SB). MLG got fundings from La Vannetaise

## Conflict of interest

The authors declare no conflict of interest.

## Author contribution

SB wrote and edited the manuscript, raised funding, designed and performed experiments, analyzed the data and generated figures, FG provided patient samples and edited the manuscript, PT analyzed IHC BCG performed the statistical analyses, EL contributed to the writing and editing of the manuscript, AP performed some of the Flow cytometry experiments, JL and VL provided patient samples. MLG supervised the work, raised funding, wrote and edited the manuscript, designed and performed experiments, analyzed the data and generated figures.

## Supplementary materials

***Supplementary Table 1- Patient and tumor characteristics.*** *For quantitative variables we give the statistical dispersion defined by the average, the standard deviation (SD), the median, the minimum and maximum (Min-Max), whereas we give percentage (%) and actual number of events (n) over total population (n total) for qualitative ones. ADD: Anxiety-depressive disorder; AHT: Arterial hypertension; BMI: Body mass index; BC: Breast carcinoma; IDC: Invasive ductal carcinoma; ILC: Invasive lobular carcinoma; DCIS: Ductal carcinoma in-situ; IV: Intra-vascular; SLN: Sentinel lymph node; LN: Lymph node; pTNM: pathological staging of tumor size(T). lymph node involvement(N). and metastatic status(M) of breast cancer according to international guidelines*.

***Supplementary Table 2- Treatment, and follow-up characteristics.*** *For quantitative variables we give the statistical dispersion defined by the average, the standard deviation (SD), the median, the minimum and maximum (Min-Max), whereas we give percentage (%) and actual number of events (n) over total population (n total) for qualitative ones. SLN: Sentinel lymph node dissection; LND: Lymph node dissection; LN: Lymph nodes. Chemotherapy regimen were mainly FEC (5-flurouracile, Epirubicine, Cyclophosphamide) and Taxanes for all but eight patients who got either Taxanes for half of them (alone or with targeted therapy), or Cyclophosphamide for the other half (+ Doxorubicine for two or Taxanes for the other two). Three of these patients also got targeted therapy by Cobimetinib for two and Olaparib for one of them*.

***Supplementary Table 3- Tumor infiltrating lymphocytes (TILs) count and immune infiltrate characterization by flow cytometry (FC).*** *For quantitative variables we give the statistical dispersion defined by the average, the standard deviation (SD), the median, the minimum and maximum (Min-Max), whereas we give percentage (%) and actual number of events (n) over total population (n total) for qualitative ones. TILs: Tumor-infiltrating lymphocytes; FC: Flow cytometry; BCSC: Breast Cancer Stem Cells; DBCC: Differentiated Breast Cancer Cells*.

***Supplementary Table 4- TILs and Th2 correlation with patient, tumor, and treatment characteristics.*** *p values are obtained thanks to a chi2 or a Fisher’s exact test (when there are less than 5 patients per group) for TILs group comparison of qualitative values and an ANOVA one for quantitative values. For TILs percentages and Th2 response comparisons, p values are obtained from a Student or an ANOVA test (when there are more than 2 groups) for qualitative values and a Pearson correlation one for quantitative values. p values in bold font are significant ones. HRT: Hormone Replacement Therapy; BMI: Body mass index; BC: Breast carcinoma; S.B.R.: Scarff-Bloom and Richardson; DCIS: Ductal carcinoma in-situ; IV: Intra-vascular; LN: Lymph Node; pTNM: pathological staging of tumor size(T), lymph node involvement (N), and metastatic status(M) of breast cancer according to international guidelines*.

***Supplementary Table 5- Correlation of patient, tumor, immunity, and treatment characteristics to relapse-free survival (RFS) or overall survival (OS).*** *p values for relapse status are obtained using Wilcoxon test for quantitative variables and Chi2 or Fisher exact test for qualitative ones (if the group does or does not contain at least 5 values respectively). p values for survival regression are obtained using log rank test for univariate (univar) analyses and Cox model for multivariate ones (multivar), using BIC condition for prognostic usefulness. p values <0.05 appear in bold font in the text. *p values/ HR [CI95%]. BMI: Body mass index; BC: Breast cancer; DCIS: Ductal carcinoma in-situ; IV: Intra-vascular; LN: Lymph Nodes; pTNM: pathological staging of tumor size (T), lymph node involvement (N), and metastatic status (M) of breast cancer according to international guidelines; TILs: Tumor-infiltrating lymphocytes; FC: Flow cytometry; BCSC: Breast Cancer Stem Cells; DBCC: Differentiated Breast Cancer Cells*.

***Supplementary Figure 1- Flow chart for patient inclusion.*** *HR+ +/− Her2+: hormonal receptors and/or Her2 positive; BC: breast cancers; TNBC: Triple-negative breast cancers; TNBC only*: exclusion of patients having developed another histological type of cancer or a recurrence as an HR+ BC; TILs: Tumor-infiltrating lymphocytes; w/o: without; w/: with; FC: Flow Cytometry. Exclusion criteria are depicted in grey*.

***Supplementary Figure 2 – Flow Cytometry (FC) characterization of tumor cells and tumor-infiltrating immune cells according to their cluster of differentiation (CD).*** *BC: Breast cancer; SC: Stem cells; DBCC: Differentiated breast cancer cells; CD4+: helper T lymphocytes; CD8+: cytotoxic T lymphocytes; BT; lab number*.

***Supplementary Figure 3- Relapse-free survival (A) and overall survival (B) for the entire cohort.*** *Kaplan Meier curves were estimated, and percentages of survival rates derived from a log-rank test*.

## Notes

### Competing Interest Statement

The authors have declared no competing interest.

### Funding Statement

This study was funded by grants from Rennes University Hospital (CORECT 2021-UF 8946-03) and association La Vannetaise

### Author Declarations

The Ethics Committee Review Board of Centre de lutte contre le Cancer Eugene Marquis gave ethical approval for this work

