## Supplementary Tables and Figures for "Th2 infiltration is a better predictor of survival than tumor-infiltrating lymphocytes (TILs) in triple-negative breast cancer (TNBC)"

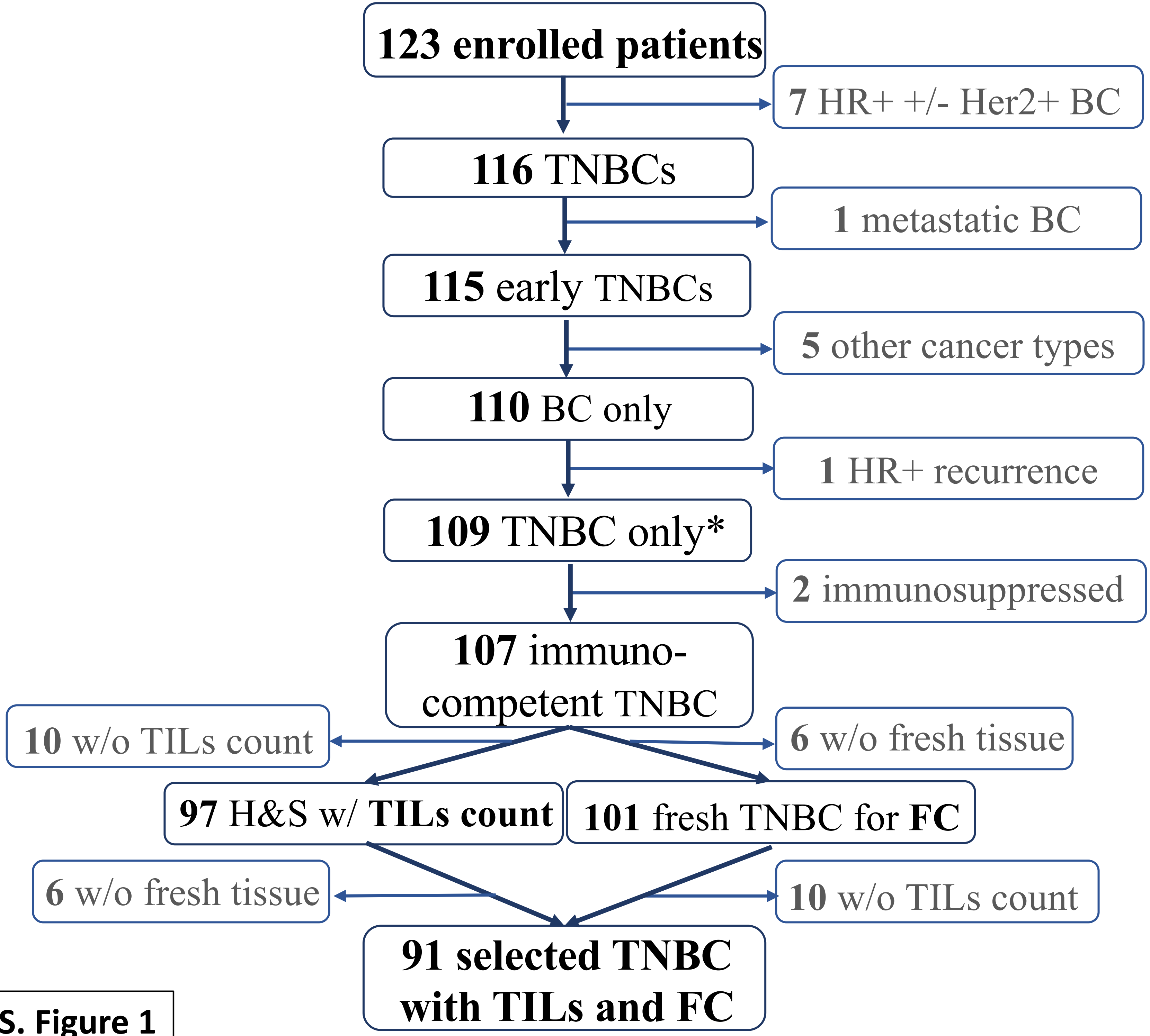
123 enrolled patients
7 HR+ +/- Her2+ BC
116 TNBCs
1 metastatic BC
115 early TNBCs
5 other cancer types
110 BC only
1 HR+ recurrence
109 TNBC only*
2 immunosuppressed
107 immuno-competent TNBC
10 w/o TILs count
6 w/o fresh tissue
97 H&S w/ TILs count
101 fresh TNBC for FC
6 w/o fresh tissue
10 w/o TILs count
91 selected TNBC with TILs and FC

S. Figure 1

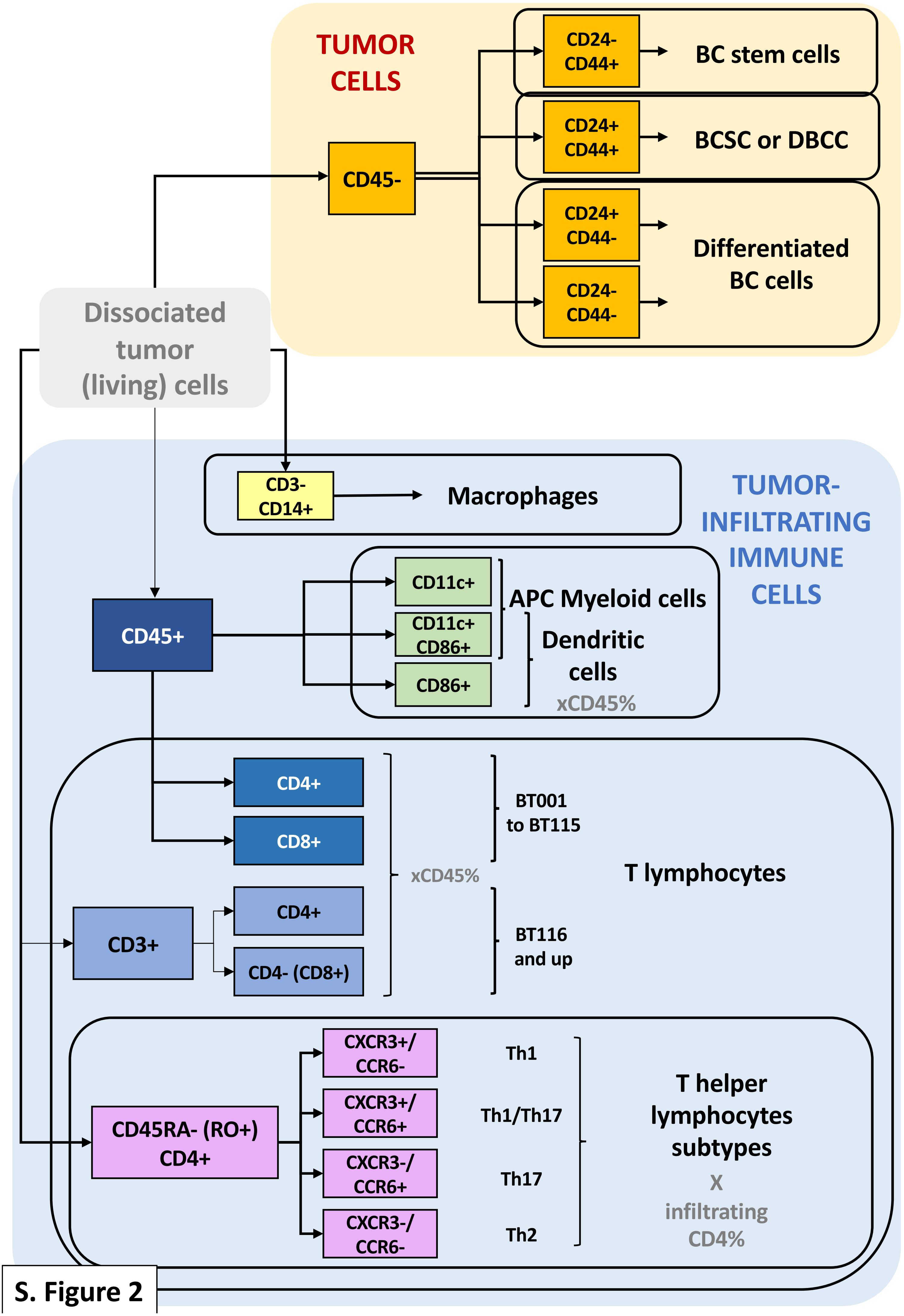
TUMOR CELLS
CD45-
CD24- CD44+
BC stem cells
CD24+ CD44+
BCSC or DBCC
CD24+ CD44-
CD24- CD44-
Differentiated BC cells
Dissociated tumor (living) cells
TUMOR-INFILTRATING IMMUNE CELLS
CD3- CD14+
Macrophages
CD45+
CD11c+
CD11c+ CD86+
CD86+
APC Myeloid cells
Dendritic cells
xCD45%
CD4+
CD8+
BT001 to BT115
xCD45%
T lymphocytes
CD3+
CD4+
CD4- (CD8+)
BT116 and up
CD45RA- (RO+) CD4+
CXCR3+/ CCR6-
Th1
CXCR3+/ CCR6+
Th1/Th17
CXCR3-/ CCR6+
Th17
CXCR3-/ CCR6-
Th2
T helper lymphocytes subtypes
X
infiltrating CD4%

S. Figure 2

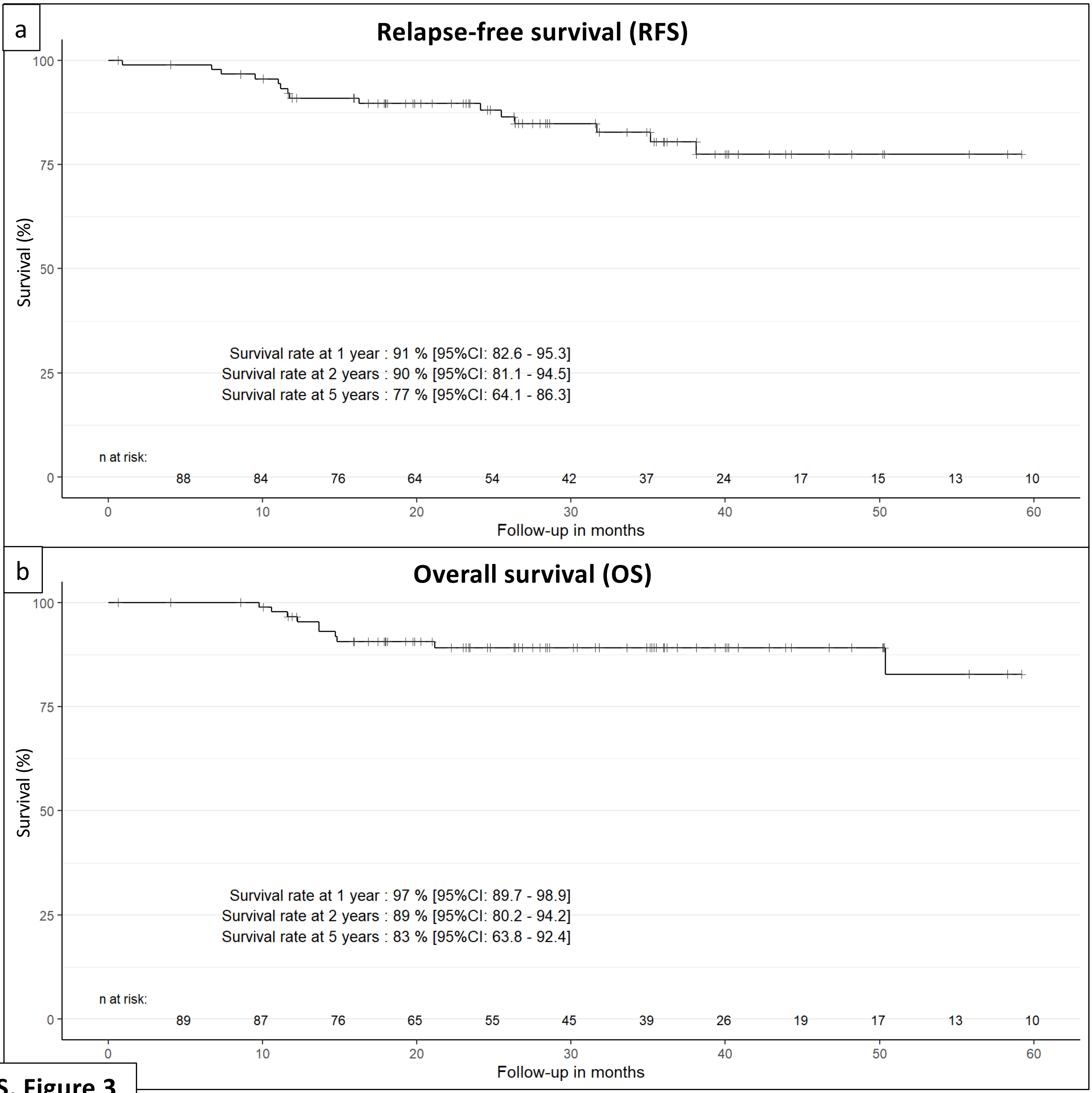
a
Relapse-free survival (RFS)
Survival (%)
100
75
50
25
0
Survival rate at 1 year : 91 % [95%CI: 82.6 - 95.3]
Survival rate at 2 years : 90 % [95%CI: 81.1 - 94.5]
Survival rate at 5 years : 77 % [95%CI: 64.1 - 86.3]
n at risk:
88 84 76 64 54 42 37 24 17 15 13 10
0 10 20 30 40 50 60
Follow-up in months
b
Overall survival (OS)
Survival (%)
100
75
50
25
0
Survival rate at 1 year : 97 % [95%CI: 89.7 - 98.9]
Survival rate at 2 years : 89 % [95%CI: 80.2 - 94.2]
Survival rate at 5 years : 83 % [95%CI: 63.8 - 92.4]
n at risk:
89 87 76 65 55 45 39 26 19 17 13 10
0 10 20 30 40 50 60
Follow-up in months

**S. Figure 3**

| Characteristics | | | Average/ % | SD/ n | Median / n total | Min-Max |
|---|---|---|---|---|---|---|
| **Patients** | Age (years) | | 58.0 | 14.5 | 57.3 | 33.4-89.1 |
| | Hormonal status | Parity : 0 | 6.7 | 6 | 90 | |
| | | 1 | 13.3 | 12 | 90 | |
| | | >1 | 80.0 | 72 | 90 | |
| | | Menopause | 59.3 | 54 | 91 | |
| | | Hormone therapy | 12.1 | 11 | 91 | |
| | Mutational profile | BRCA1 | 15.7 | 8 | 51 | |
| | | BRCA2 | 11.8 | 6 | 51 | |
| | | Other mutations | 0.0 | 0 | 46 | |
| | Personal medical history | Breast cancer | 13.2 | 12 | 91 | |
| | | Ovarian cancer | 0.0 | 0 | 91 | |
| | | Comorbidities : None | 50.5 | 46 | 91 | |
| | | Dysthyroidism | 14.3 | 13 | 91 | |
| | | Diabetes | 1.1 | 1 | 91 | |
| | | Asthma | 6.6 | 6 | 91 | |
| | | ADD | 8.8 | 8 | | |
| | | AHT | 19.8 | 18 | 91 | |
| | | BMI (kg/m²) | 24.7 | 4.5 | 24.8 | 14.0-40.1 |
| | Familial history | Breast cancer | 39.6 | 36 | 91 | |
| | | Ovarian cancer | 7.7 | 7 | 91 | |
| **Tumor** | Side | Right | 48.4 | 44 | 91 | |
| | | Left | 51.6 | 47 | 91 | |
| | Foci | One | 82.2 | 74 | 90 | |
| | | Multiple | 17.8 | 16 | 90 | |
| | Hystological type of BC | IDC | 85.7 | 78 | 91 | |
| | | ILC | 2.2 | 2 | 91 | |
| | | Medullary | 1.1 | 1 | 91 | |
| | | Metaplastic | 1.1 | 1 | 91 | |
| | | Other | 9.9 | 9 | 91 | |
| | Grade (Surgery) | 1 | 1.1 | 1 | 91 | |
| | | 2 | 5.5 | 5 | 91 | |
| | | 3 | 93.4 | 85 | 91 | |
| | | Ki67 (%) | 64.8 | 23.8 | 70.0 | 5.0-95.0 |
| | DCIS | Presence | 44.9 | 40 | 89 | |
| | | Size (mm) | 18.8 | 12.9 | 22.5 | 2.0-45.0 |
| | | Grade : Unknown | 5.0 | 2 | 40 | |
| | | 2 | 2.5 | 1 | 40 | |
| | | 3 | 92.5 | 37 | 40 | |
| | Size (mm) | Mammographical | 21.9 | 10.0 | 20.0 | 7.0-70.0 |
| | | Pathological | 22.9 | 11.6 | 20.0 | 8.0-85.0 |
| | Presence of IV tumor embol | | 14.6 | 13 | 89 | |
| | Number of SLN | Harvested | 1.4 | 1.3 | 1.0 | 0.0-5.0 |
| | | Metastatic | 0.3 | 0.6 | 0.0 | 0.0-2.0 |
| | Total number of LN | Harvested | 6.2 | 6.1 | 4.0 | 0.0-26.0 |
| | | Metastatic | 1.5 | 3.6 | 0.0 | 0.0-21.0 |
| | pTNM | Size : T1 | 45.1 | 41 | 91 | |
| | | T2 | 51.6 | 47 | 91 | |
| | | T3 | 2.2 | 2 | 91 | |
| | | T4 | 1.1 | 1 | 91 | |
| | | LN involvement : N0 | 62.6 | 57 | 91 | |
| | | N1 | 22.0 | 20 | 91 | |
| | | N2 | 7.7 | 7 | 91 | |
| | | N3 | 3.3 | 3 | 91 | |
| | | Nx | 4.4 | 4 | 91 | |
| | | Metastasis : M0 | 96.7 | 88 | 91 | |
| | | M1 | 1.1 | 1 | 91 | |
| | | Mx | 2.2 | 2 | 91 | |

***Supplementary Table 1- Patient and tumor characteristics.*** *For quantitative variables we give the statistical dispersion defined by the average, the standard deviation (SD), the median, the minimum and maximum (Min-Max), whereas we give percentage (%) and actual number of events (n) over total population (n total) for qualitative ones. ADD: Anxiety-depressive disorder; AHT: Arterial hypertension; BMI: Body mass index; BC: Breast carcinoma; IDC: Invasive ductal carcinoma; ILC: Invasive lobular carcinoma; DCIS: Ductal carcinoma in-situ; IV: Intra-vascular; SLN: Sentinel lymph node; LN: Lymph node; pTNM: pathological staging of tumor size(T). lymph node involvement(N). and metastatic status(M) of breast cancer according to international guidelines.*

| Characteristics | | | Average/ % | SD/ n | Median / n total | Min-Max |
|---|---|---|---|---|---|---|
| ***Treatment*** | Neoadjuvant therapy | Chemotherapy | 1.1 | 1 | 91 | |
| | | Other therapy | 0.0 | 0 | 91 | |
| | Surgery | Breast: Lumpectomy | 68.1 | 62 | 91 | |
| | | Breast-conserving | 7.7 | 7 | 91 | |
| | | Mastectomy | 24.2 | 22 | 91 | |
| | | Axilla: SLN | 54.4 | 49 | 90 | |
| | | LND | 28.9 | 26 | 90 | |
| | | SLN+LND | 11.1 | 10 | 90 | |
| | | No surgery | 5.6 | 5 | 90 | |
| | Adjuvant therapy | Chemotherapy | 85.7 | 78 | 91 | |
| | | Targeted Therapy | 3.4 | 3 | 88 | |
| | | Radiotherapy: Breast | 86.5 | 77 | 89 | |
| | | Local Boost | 62.3 | 43 | 69 | |
| | | LN | 38.0 | 27 | 71 | |
| ***Follow-up*** | Time (months) | All patients | 32.3 | 17.9 | 28.0 | 0.0-76.0 |
| | | Alive ones | 34.2 | 17.6 | 31.0 | 0.0-76.0 |
| | Locoregional recurrence | Presence | 7.7 | 7 | 91 | |
| | | Time to (months) | 24.6 | 23.0 | 24.0 | 6.0-73.0 |
| | Metastases | Bone | 2.2 | 2 | 91 | |
| | | Nervous System | 6.6 | 6 | 91 | |
| | | Other visceral ones: All | 12.1 | 11 | 91 | |
| | | Lung | 81.8 | 9 | 11 | |
| | | Liver | 45.5 | 5 | 11 | |
| | | Other sites | 18.2 | 2 | 11 | |
| | | Time to (months) | 17.2 | 11.6 | 11.0 | 0.0-38.0 |
| | All kinds of relapses | % | 17.6 | 16 | 91 | |
| | | Time to (months) | 20.9 | 17.8 | 13.5 | 0.0-73.0 |
| | Death due to cancer | % | 11.0 | 10 | 91 | |
| | | Time to (months) | 16.7 | 12.1 | 13.0 | 9.0-50.0 |

***Supplementary Table 2- Treatment, and follow-up characteristics.*** *For quantitative variables we give the statistical dispersion defined by the average, the standard deviation (SD), the median, the minimum and maximum (Min-Max), whereas we give percentage (%) and actual number of events (n) over total population (n total) for qualitative ones. SLN : Sentinel lymph node dissection ; LND : Lymph node dissection; LN : Lymph nodes. Chemotherapy regimen were mainly FEC (5-flurouracile, Epirubicine, Cyclophosphamide) and Taxanes for all but eight patients who got either Taxanes for half of them (alone or with targeted therapy), or Cyclophosphamide for the other half (+ Doxorubicine for two or Taxanes for the other two). Three of these patients also got targeted therapy by Cobimetinib for two and Olaparib for one of them.*

| Characteristics | | *Average/ %* | *SD/ n* | *Median / n total* | *Min-Max* |
|---|---|---|---|---|---|
| ***TILs*** | Group: A | 58.2 | 53 | 91 | |
| | B | 23.1 | 21 | 91 | |
| | C | 18.7 | 17 | 91 | |
| | % | 22.8 | 25.9 | 10.0 | 0.0-90.0 |
| ***FC Characterization*** | Tumor cells | 72.1 | 25.8 | 80.0 | 3.5-98.5 |
| | BCSC/DBCC (CD44+CD24+) | 5.5 | 8.8 | 1.1 | 0.0-42.3 |
| | BCSC (CD44+CD24-) | 10.0 | 17.9 | 2.6 | 0.0-86.8 |
| | DBCC (CD44-CD24+) | 15.8 | 22.4 | 4.8 | 0.0-88.5 |
| | DBCC (CD44-CD24-) | 68.6 | 33.3 | 84.8 | 5.1-99.9 |
| | Total leucocytes | 36.0 | 30.4 | 22.9 | 1.4-96.0 |
| | Helper T cells | 14.1 | 13.1 | 9.7 | 0.0-59.4 |
| | Cytotoxic T cells | 11.1 | 10.8 | 8.1 | 0.0-52.2 |
| | Th1 response | 1.0 | 1.7 | 0.3 | 0.0-7.2 |
| | Th1/Th17 response | 0.5 | 1.1 | 0.1 | 0.0-6.8 |
| | Th17 response | 0.8 | 2.2 | 0.1 | 0.0-18.8 |
| | Th2 response | 11.7 | 12.1 | 8.0 | 0.0-59.0 |
| | Th1/Th2 response | 0.7 | 4.4 | 0.1 | 0.0-41.5 |
| | Myeloid cells (CD11c+) | 8.4 | 16.6 | 1.3 | 0.0-70.2 |
| | Dendritic cells (CD11c+86+) | 2.1 | 9.6 | 0.1 | 0.0-66.1 |
| | Dendritic cells (CD86+) | 1.0 | 2.2 | 0.2 | 0.0-10.3 |
| | Macrophages | 3.1 | 6.2 | 0.9 | 0.1-32.2 |

***Supplementary Table 3- Tumor infiltrating lymphocytes (TILs) and flow cytometry (FC) characteristics.*** *For quantitative variables we give the statistical dispersion defined by the average, the standard deviation (SD), the median, the minimum and maximum (Min-Max), whereas we give percentage (%) and actual number of events (n) over total population (n total) for qualitative ones. TILs: Tumor-infiltrating lymphocytes; FC: Flow cytometry; BCSC: Breast Cancer Stem Cells; DBCC: Differentiated Breast Cancer Cells.*

| Characteristics | | | TILs | | Th response | |
|---|---|---|---|---|---|---|
| | | | *Group* | *%* | *Th2* | *Th1/Th2* |
| ***Patients*** | Age (years) | | 0.927 | 0.567 | 0.667 | 0.170 |
| | Hormonal status | Parity | 0.357 | 0.301 | 0.609 | 0.798 |
| | | Menopause | 0.981 | 0.784 | 1 | 0.384 |
| | | HRT | 0.819 | 0.355 | 0.545 | 0.203 |
| | BRCA Mutation | | **0.009** | **0.016** | 0.478 | 0.416 |
| | Personal history | Breast cancer | 0.762 | 0.560 | 0.929 | 0.272 |
| | | Ovarian cancer | - | - | - | - |
| | | BMI (kg/m²) | **0.038** | **0.007** | 0.732 | 0.866 |
| | Familial history | Breast cancer | 0.835 | 0.899 | 0.966 | 0.257 |
| | | Ovarian cancer | **0.018** | 0.191 | 0.387 | 0.302 |
| ***Tumor*** | Side | | 0.915 | 0.846 | 0.604 | 0.240 |
| | Uni- or Multifocal | | 0.472 | 0.301 | 0.171 | 0.328 |
| | Histological type of BC | | 0.754 | - | - | - |
| | S.B.R. Grade | | 0.503 | **1.78.10$^{-9}$** | 0.180 | 0.206 |
| | Ki67 (%) | | 0.325 | 0.169 | 0.763 | 0.198 |
| | Presence of DCIS | | **0.049** | **0.037** | 0.562 | 0.414 |
| | Presence of IV tumor emboli | | 0.453 | 0.601 | **0.047** | 0.284 |
| | Size (mm) | Pathological | 0.689 | 0.637 | 0.979 | 0.835 |
| | Number of metastatic LN | | 0.325 | 0.283 | 0.609 | 0.634 |
| | pTNM | T(Size) | 0.704 | 0.801 | 0.209 | |
| | | N (LN) | 0.410 | 0.998 | 0.123 | |
| | | M (Metastasis) | 0.191 | - | - | - |
| ***Treatment*** | Surgery | Breast | 0.992 | 0.828 | 0.278 | |
| | | Axillary | 0.584 | 0.320 | 0.392 | |

***Supplementary Table 4- TILs and Th2 correlation with patient, tumor, and treatment characteristics.*** *p values are obtained thanks to a chi2 or a Fisher's exact test (when there are less than 5 patients per group) for TILs group comparison of qualitative values and an ANOVA one for quantitative values. For TILs percentages and Th2 response comparisons, p values are obtained from a Student or an ANOVA test (when there are more than 2 groups) for qualitative values and a Pearson correlation one for quantitative values. p values in bold font are significant ones. HRT : Hormone Replacement Therapy; BMI: Body mass index; BC: Breast carcinoma; S.B.R.: Scarff-Bloom and Richardson; DCIS: Ductal carcinoma in-situ; IV: Intra-vascular; LN: Lymph Node; pTNM: pathological staging of tumor size(T). lymph node involvement(N). and metastatic status(M) of breast cancer according to international guidelines.*

| **Characteristics** | | | ***Correlation to RFS (p)*** | | | ***Correlation to OS (p)*** | |
|---|---|---|---|---|---|---|---|
| | | | *Relapse Status* | *Survival* | | *Living Status Survival* | |
| | | | | *Univar* | *Multivar* | *Univar* | *Multivar* |
| ***Patients*** | Age (years) | | 0.538 | 0.762 | | 0.292 | |
| | Hormonal status | Parity | 0.337 | 0.076 | | 0.478 | |
| | | Menopause | 0.735 | 0.974 | | 0.625 | |
| | | Hormone therapy | 1 | 0.160 | | 0.942 | |
| | Mutational profile | BRCA1 | 0.234 | 0.168 | | 0.160 | |
| | | BRCA2 | 0.421 | 0.308 | | 0.279 | |
| | | Other mutations | NA | | | NA | |
| | Personal medical history | Breast cancer | 0.348 | 0.226 | | 0.192 | |
| | | Ovarian cancer | NA | | | NA | |
| | | BMI | **0.033** | 0.205 | | **0.034** | |
| | Familial medical history | Breast cancer | 1 | 0.481 | | 0.919 | |
| | | Ovarian cancer | 0.735 | 0.257 | | 0.359 | |
| ***Tumor*** | Side | | 0.741 | 0.412 | | 0.616 | |
| | One or multiple foci | | **0.017** | 0.111 | | **0.009** | |
| | Histological type of BC | | 0.549 | 0.719 | | 0.693 | |
| | Grade (Surgery) | | 0.513 | 0.905 | | 0.783 | |
| | Ki67 (%) | | 0.568 | 0.719 | | 0.554 | |
| | DCIS | Presence | 0.289 | 0.796 | | 0.278 | |
| | | Size (mm) | 0.127 | 0.317 | | 0.317 | |
| | | Grade | 0.383 | 0.842 | | 0.544 | |
| | Pathological size (mm) | | **0.002** | 0.064 | **0.011/ 0.23[0.07-0.78]*** | **0.010** | **0.013/, 0.13[0.02-1.01]*** |
| | IV tumor emboli | | 0.122 | 0.363 | | 0.096 | |
| | Total number of LN | Harvested | 0.280 | 0.433 | | 0.368 | |
| | | Metastatic | **0.014** | **0.004** | | 0.068 | |
| | pTNM | T | 0.233 | 0.440 | | 0.252 | |
| | | N | **0.036** | **0.002** | **0.004/ 2.85[2.35-26.25]*** | **0.024** | **0.016/ 5.67[1.26-25.42]*** |
| | | M | **0.011** | **<0.0001** | | **<0.0001** | |
| ***Immunity*** | TILs | Group | 0.102 | 0.164 | | 0.121 | |
| | | % | 0.102 | 0.071 | | **0.044** | |
| | FC characterization | Tumor cells | 0.284 | 0.190 | | 0.338 | |
| | | BCSC (CD44+CD24+) | 0.944 | 0.168 | | 0.421 | |
| | | BCSC (CD44+CD24-) | 0.899 | 0.821 | | 0.597 | |
| | | DBCC (CD44-CD24+) | 0.889 | 0.710 | | 0.802 | |
| | | DBCC (CD44-CD24-) | 0.603 | 0.324 | | 0.385 | |
| | | Total lymphocytes | 0.319 | 0.243 | | 0.377 | |
| | | Helper T cells | 0.406 | 0.306 | | 0.432 | |
| | | Cytotoxic T cells | 0.153 | 0.072 | | 0.188 | |
| | | Th1 response | 0.864 | 0.245 | | 0.474 | |
| | | Th1/Th17 response | 0.894 | 0.642 | | 0.390 | |
| | | Th17 response | 0.643 | 0.476 | | 0.564 | |
| | | Th2 response | 0.177 | **0.009** | **0.002/ 9.30[2.38-36.26]*** | **0.008** | **0.010/ 11.46[1.36-96.39]*** |
| | | Myeloid cells (CD11c+) | **0.012** | 0.265 | | 0.182 | |
| | | Dendritic cells (CD11c+CD86+) | **0.030** | **0.030** | | 0.259 | |
| | | Dendritic cells (CD86+) | 0.061 | **0.028** | | 0.250 | |
| | | Macrophages | 0.653 | 0.714 | | 0.660 | |
| ***Treatment*** | Surgery | Breast | 0.284 | 0.435 | | 0.220 | |
| | | Axilla | 0.167 | 0.564 | | 0.404 | |
| | Adjuvant therapy | Chemotherapy | 0.631 | 0.441 | | 0.517 | |
| | | Targeted Therapy | 0.307 | 0.284 | | 0.138 | |
| | | Radiotherapy: Breast | 0.619 | 0.593 | | 0.610 | |
| | | Local Boost | **0.027** | 0.146 | | 0.062 | |
| | | LN | 0.489 | 0.274 | | 0.143 | |

***Supplementary Table 5- Correlation of patient, tumor, treatment, and follow-up characteristics to relapse-free survival (RFS) or overall survival (OS).*** *p values for relapse status are obtained using Wilcoxon test for quantitative variables and Chi2 or Fisher exact test for qualitative ones, depending if the group do or do not contain at least 5 values respectively. p values for survival regression are obtained using log rank test for univariate (univar) analyses and Cox model for multivariate ones (multivar), using BIC condition for prognostic usefulness. p values <0.05 appear in bold font in the text. *p values/ HR [CI95%]. BMI : Body mass index ; BC: Breast cancer ; DCIS : Ductal carcinoma in-situ ; IV : Intra-vascular ; LN : Lymph Nodes ; TILs : Tumor-infiltrating lymphocytes ; FC : Flow cytometry ; BCSC : Breast Cancer Stem Cells ; DBCC : Differentiated Breast Cancer Cells ; pTNM : pathological staging of tumor size(T), lymph node involvement(N), and metastatic status(M) of breast cancer according to international guidelines.*
